# Towards Culture-Free Sequencing of *Mycobacterium tuberculosis*: Evaluating New Targeted and Whole-Genome Approaches for Genotyping and Drug Resistance Profiling

**DOI:** 10.64898/2026.01.15.26344185

**Authors:** Ilaria Iannucci, Federico Di Marco, Kiarash Moghaddasi, Paolo Miotto, POR TB study group, Andrea Maurizio Cabibbe, Daniela Maria Cirillo

## Abstract

**Background:** The effectiveness of current drug-resistant tuberculosis (DR-TB) regimens is limited by the absence of rapid diagnostics that comprehensively predict resistance to included drugs. Next-generation sequencing (NGS), through culture-free targeted sequencing (tNGS) and culture-based whole-genome sequencing (cWGS) of the *Mycobacterium tuberculosis* complex (MTBC), offers a powerful framework for precision diagnosis, surveillance, and trial applications. We evaluated two novel assays enabling high-resolution tNGS and enrichment-based direct WGS (dWGS) on respiratory samples, focusing on analytical sensitivity, DR prediction accuracy, and genotyping concordance.

**Methods:** tNGS Deeplex Myc-TB XL tNGS (Genoscreen, beta-testing) and dWGS QIAseq xHYB MTB (Qiagen) were evaluated on 96 MTBC–positive decontaminated sputum samples from a vaccine trial, spanning a wide range of bacillary loads. DNA was extracted using a host-depletion protocol and quantified by MTBC-specific real-time PCR. Libraries were sequenced on Illumina platforms and analysed using assay-specific pipelines. Associations between genome copy (gc) number and sequencing coverage were assessed. DR prediction performance was benchmarked against cWGS and the WHO mutation catalogue across first-, second-line, newer, and repurposed drugs. WGS-based phylogenetic trees were constructed using Ridom SeqSphere+.

**Results:** Bacillary loads ranged from <10 to >1,000 MTBC gc/µL. tNGS generated interpretable resistance profiles in 96.6% of specimens, achieving a limit of detection (LoD) of ∼10 gc, with 100% sensitivity for all evaluated drugs and 100% specificity except for isoniazid/ethionamide (≥97%). dWGS yielded data suitable for DR analysis in 75% of samples, with a LoD of ∼100gc. Sensitivity was 100% for most drugs; one fluoroquinolone-resistant case was missed due to low-frequency variant thresholds, and resistance to delamanid and clofazimine was misclassified in one case each owing to interpretation rules. Specificity was 100% except for rifampicin (≥97%). Lineage assignment was concordant with cWGS for both approaches, and dWGS enabled transmission analysis in 65% of samples, confirming the cluster detected by cWGS.

**Conclusions:** tNGS provides sensitive and specific DR profiling with a LoD comparable to the most sensitive rapid assays. dWGS, while requiring higher DNA input, enables robust culture-free genome-wide analysis, including transmission inference and exploration of candidate DR loci. Bacillary load–guided integration of both approaches may optimize DR-TB clinical management and genomic surveillance.

## Background

The continued emergence and spread of drug-resistant tuberculosis (DR-TB) pose a critical and persistent threat to TB control worldwide(1). Although WHO has recently endorsed shorter, highly effective all-oral regimens to treat rifampicin-resistant/multidrug-resistant (RR/MDR) TB, such as the 6-month BPaLM (bedaquiline, pretomanid, linezolid, moxifloxacin), the diagnostic capacity required to support individualized therapy remains inadequate(2, 3). In 2024, only 57% of people with RR-TB received fluoroquinolone susceptibility testing, while access to testing for bedaquiline and other new drugs was even more limited, both globally (1) and within the European region (4). This diagnostic shortfall represents a major barrier to effective regimen selection, stewardship of novel agents, and control of transmission. There is an urgent need for rapid, comprehensive, and scalable diagnostic tools capable of accurately guiding tailored DR-TB treatment and strengthening public health responses (4). Additionally, clinical trials for new regimens or vaccine candidates require rapid tools to properly enrol participants, to monitor emerging drug resistance determinants, and defining phylogenetic relationships to evaluate the generalizability and impact of proposed interventions.

Over the past decade, next-generation sequencing (NGS) has revolutionized TB diagnostics, surveillance and clinical trials by providing molecular characterization of resistance-associated mutations and high-resolution genotyping of the *M. tuberculosis* complex (MTBC)(5–9). Among NGS approaches, targeted sequencing (tNGS) and whole-genome sequencing (WGS) are the most common strategies for MTBC genomic characterization.

tNGS assays amplify predefined drug resistance-associated genomic regions and sequence them in parallel, allowing rapid culture-free identification of MTBC and detection of first- and second-line resistance directly from clinical specimens (6, 7). However, because of its reliance on predefined targets, its genomic coverage is inherently limited, and mutations outside the selected regions (including rare or newly emerging variants) may remain undetected. In addition, tNGS does not support transmission analysis (10) and its performance is affected by the initial bacterial load.

Among the tNGS tools recently endorsed by WHO(6, 7), the Deeplex Myc-TB (GenoScreen) is approved for detection of resistance to isoniazid (INH), pyrazinamide (PZA), ethambutol (EMB), fluoroquinolones (FQ), bedaquiline (BDQ), linezolid (LZD), clofazimine (CFZ), amikacin (AMK) and streptomycin (STM) among people with RR-TB(9, 11–13). The newer version of the assay, Deeplex Myc-TB XL, currently in beta testing, expands coverage to 29 gene targets including for new key drugs such as pretomanid (PMD), thus aligning with the updated DR-TB treatment guidelines, and aims to improve sensitivity(14).

In contrast, WGS provides comprehensive genome-wide data, enabling detection of both known and novel resistance-associated variants, assignment of MTBC lineages, and high-resolution transmission analysis. WGS, currently performed from isolates obtained in liquid or solid cultures, is considered reference standard for testing of RIF, EMB and PZA resistance, and complements phenotypic drug susceptibility test (pDST) for drug resistance interpretation(6, 7). Nonetheless, its reliance on culture introduces delays of weeks to months and requires substantial laboratory and bioinformatics infrastructure (5–7, 9). To overcome these limitations, direct WGS (dWGS) has emerged as a promising fast approach, enabling rapid full-genome analysis for resistance prediction, lineage assignment, and high-resolution genotyping directly from clinical specimens (15, 16).

Several enrichment strategies, including host DNA depletion (17) and direct capture using biotinylated probes (17–21), have been applied to enable MTBC dWGS from clinical specimens, which typically contain high background of human and non-mycobacterial DNA. These studies (17–21), demonstrated the diagnostic potential of culture-free WGS for MTBC, but still suboptimal performance.

The QIAseq xHYB *Mycobacterium tuberculosis* (MTB) Panel (Qiagen) was developed to overcome existing challenges by enabling hybrid capture and targeted enrichment of entire MTBC genome directly from clinical specimens, enabling deep full-genomic sequencing without the need of culture(22).

Despite increasing interest in both tNGS and dWGS approaches, comparative evaluations of their performance on clinical samples remain limited(15, 16). This study provides the first head-to-head comparison of an expanded tNGS assay (the new Deeplex Myc-TB XL) and an enrichment-based culture-free WGS workflow (the QIAseq xHYB MTB) on the same MTBC-positive sputum sediments(23). We assessed the limit of detection, drug resistance detection accuracy compared with culture-based WGS (cWGS) from the same samples, and for dWGS, phylogenetic clustering concordance (compared with cWGS).

## Methods

### MTBC sediments and cWGS

We analysed 96 MTBC-positive sputum sediments collected at baseline within a vaccine trial conducted in South Africa and Tanzania and designed to assess recurrence prevention (NCT03512249)(23). This randomized, double-blind, placebo-controlled phase 2b trial enrolled adults with Xpert MTB/RIF Ultra (Cepheid)-positive TB who received the study intervention after completing TB treatment. The trial demonstrated that post-treatment vaccination with H56:IC31was safe and immunogenic but did not reduce the risk of recurrent TB(23).

The samples were selected from the archived trial biobank across participating sites through a combination of random sampling and targeted inclusion of specimens exhibiting DR profiles based on cWGS analysis. All sediments were decontaminated using the NAC-PAC RED kit (Alpha Tech). MTBC culture, and cWGS were performed previously and are described in detail elsewhere(23). cWGS data were available for all corresponding isolates. Raw FASTQ files were deposited in the NCBI Sequence Read Archive under the study accession number PRJNA1171160 (23, 24). For this study, bioinformatic interpretation of resistance-associated variants was repeated using the latest WHO mutation catalogue version (14).

### DNA extraction and quantification

To maximize MTBC DNA recovery and sequencing performance, all sputum sediments underwent DNA extraction using the Ultra-Deep Microbiome Prep kit (Molzym GmbH & Co. KG) (25) which includes human host DNA depletion. Starting from 500uL of initial sample volume, extracted microbial DNA was eluted in 100µL of nuclease free water. Total extracted DNA was quantified with the Qubit dsDNA High Sensitivity assay (Thermo Fisher Scientific). To estimate the proportion of mycobacterial DNA in the extracted samples, MTBC-specific copy number was quantified using an in-house real-time PCR (qPCR) assay targeting the *senX3-regX3* locus(26).

### Deeplex Myc-TB XL library preparation, sequencing and data analysis

Compared to the Deeplex Myc-TB assay, the XL version includes additional targets: *atpE, pepQ, mmpS5, mmpl5, ddn, fbiA, fbiB, fbiC, fbiD*, and *fgd*, while *tylA* is no longer part of the panel. After amplification with the Deeplex Myc-TB XL kit as per manufacturer’s instructions, amplicon libraries were prepared using the Illumina DNA Prep Kit and sequenced with 150-bp paired-end reads on a MiniSeq high-output platform (Illumina, San Diego, CA, USA) (2 sequencing runs with 47 tests each). The procedure was conducted in line with the protocol previously applied for the standard Deeplex Myc-TB kit (12, 13), with the main modification being that the DNA clean-up step after PCR amplification is omitted in the XL protocol. Bioinformatic analyses were performed using the dedicated proprietary bioinformatics pipeline (beta testing version), which refers to the lasted WHO MTBC mutation catalogue (14) for drug resistance interpretation. tNGS raw FASTQ files generated in this study were deposited in the NCBI Sequence Read Archive under study accession number PRJNA1392017 (27).

### QIAseq xHYB MTB library preparation, sequencing and data analysis

Libraries for dWGS were prepared with the QIAseq xHYB MTB Library Kit (Qiagen, ID. 334502 version 07/2024)(22), which enables targeted whole genome MTBC enrichment by tiling hybrid capture probes (**Additional file 4 - Suppl. Fig. 2B**).

DNA extracts were enzymatically fragmented and converted into Illumina-compatible libraries with the QIAseq FX DNA Library Kit and Unique Dual Indexes. After end repair and A-tailing, sequencing adapters were ligated, libraries were purified with QIAseq Beads to remove free adapters and subsequently amplified to obtain sufficient material for hybrid capture. For targeted enrichment, amplified libraries were pooled according to qPCR quantification to balance bacterial load across samples. Hybrid capture was performed using the QIAseq xHYB MTB panel, which employs tiled biotinylated probes across the TB genome. Libraries and probes were denatured, hybridized overnight, and captured on streptavidin-coated beads. After washing to remove non-specific products, bound libraries were eluted, re-amplified, and purified with QIAseq Beads to generate final double-stranded sequencing libraries.

Sequencing was carried out on a NextSeq 500/550 mid-output platform (Illumina) with 150-bp paired-end reads (2 runs, 48 samples each). Bioinformatic analyses were performed using CLC Genomics Workbench version 25.0.2 (Qiagen), which provides quality control, read processing, and variant calling. Drug resistance predictions were generated based on the most recent WHO Catalogue of Mutations for MTBC (14). Resistance was inferred using predefined allele-frequency thresholds. A cutoff of ≥80% was applied specifically to the *rpoB* Ser441Ala variant and to *rrs* mutations associated with AMK, KAN, and CAP resistance, to mitigate potential single nucleotide polymorphism (SNP)-calling noise attributable to low-level contamination by closely related bacterial species. For all other resistance-associated mutations, a threshold of 20% was used. Mutation analysis was restricted to samples with mean genome depth greater than or equal to 20x and complete genome breadth. dWGS raw FASTQ files generated in this study were deposited in the NCBI Sequence Read Archive under study accession number PRJNA1392017 (27).

### Statistical analysis

Comparison between estimated number of genome copies (gc) (expressed as Cycle thresholds (Ct) from qPCR) and genome coverage from dWGS or gene target coverages from tNGS were conducted performing linear regression and reporting the R^2^ score. All data processing and statistical computations were conducted within the R statistical computing environment, version 4.1.2 (28).

The number of study samples included in the performance assessment (success vs. failure) and DR/genotyping analyses for each approach is shown in **Additional file 4 - Suppl. Fig. 2A.**

We calculated sensitivity and specificity to evaluate the agreement between cWGS and the QIAseq xHYB MTB or Deeplex Myc-TB XL kit results for drug resistance classification. The following drugs were evaluated: AMK, Kanamycin (KAN), Capreomycin (CAP), BDQ, CFZ, delamanid (DLM), PMD, ethionamide (ETH), INH, EMB, levofloxacin (LEV), moxifloxacin (MOX), LZD, PZA, RIF, STM. A binary outcome classification (1=Resistant, 0=Sensitive) was applied for each platform and drug.

A confusion matrix was constructed for each drug to determine True Positives (TP), False Negatives (FN), True Negatives (TN), and False Positives (FP). Sensitivity and Specificity were calculated using the standard formulas: Sensitivity = TP / (TP + FN) * 100; Specificity = TN / (TN + FP) * 100; while confidence intervals at 95% were computed using the exact binomial test.

Only drugs for which the reference standard detected at least one resistant sample (TP + FN > 0) were included in the results.

### Transmission analysis

Transmission analysis was conducted on MTBC samples with suitable cWGS and dWGS data quality. A predefined core genome multi-locus sequence typing (cgMLST) scheme implemented in Ridom SeqSphere+ software v.10.0.5 (Ridom GmbH) was used to assess genetic relationships between isolates, excluding samples with more than 10% missing values in distance columns. Minimum spanning trees (MSTs) were generated to visualize relatedness, and a cut-off of ≤5 allelic differences was applied to define closely related strains (clusters) (5).

## Results

### Quality metrics

Quantification of MTBC gc by qPCR revealed a wide distribution across samples, ranging from <10 gc/µL (Ct 31–37; mean 2.6 gc/µL; mean Ct 33.9) to >1000 gc/µL (Ct <24; mean 4012 gc/µL; mean Ct 23.2) (**Additional file 4 - Suppl. Fig. 1A–B; Additional file 1**).

Illumina sequencing was successfully completed for 89/91 samples plus 3/3 controls evaluated with the Deeplex Myc-TB XL tNGS assay and for all 96/96 samples processed with the QIAseq xHYB MTB dWGS assay (**Additional file 4 - Suppl. Fig. 2A**). Two samples were excluded from Deeplex Myc-TB XL analysis for technical issues during library preparation.

Interpretable MTBC sequencing profiles were obtained for 86/89 study samples (96.6%; mean Ct = 28.4 ± 3.2) processed with the Deeplex Myc-TB XL assay, and for the three controls used. Sequencing results for three study samples were of insufficient quality and could not be analysed due to lower coverage depth (<200×) and breadth (<75%): mean Ct 34.2 ± 0.8 (**Fig. 1; Additional file 1**). These three samples had only 5/29, 15/29, and 2/29 assay targets covered by reads, respectively, whereas the 86 analysable samples showed ≥19/29 targets fully covered by reads.

**Figure 1:**
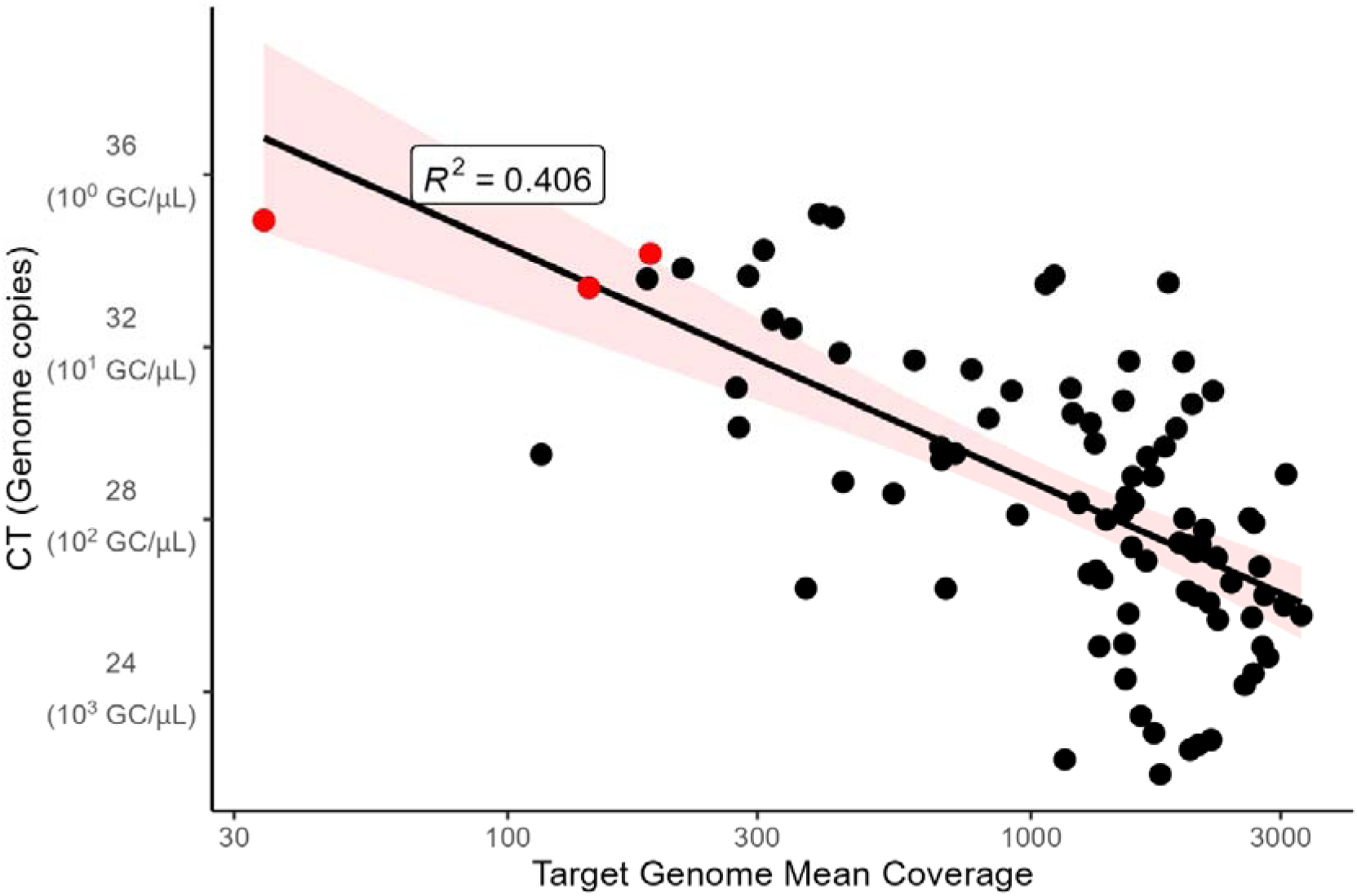
Correlation correlation between Deeplex Myc-TB XL data mean depth coverage (in 29 target regions) and Ct values for 89 MTBC sediments analyzed with tNGS. The coefficient of determination (R^2^) represents the proportion of variance from the linear interpolation.

Samples processed with the QIAseq xHYB MTB assay yielded high-quality MTBC data (depth >20×, breadth >98%) for 72/96 samples (75%; mean Ct 27.9±2.8), enabling drug resistance analysis. The remaining 24 samples (25.0%; mean 31.6 ±4.2) produced incomplete genome data (depth <20×) (**Fig. 2; Additional file 1**).

**Figure 2:**
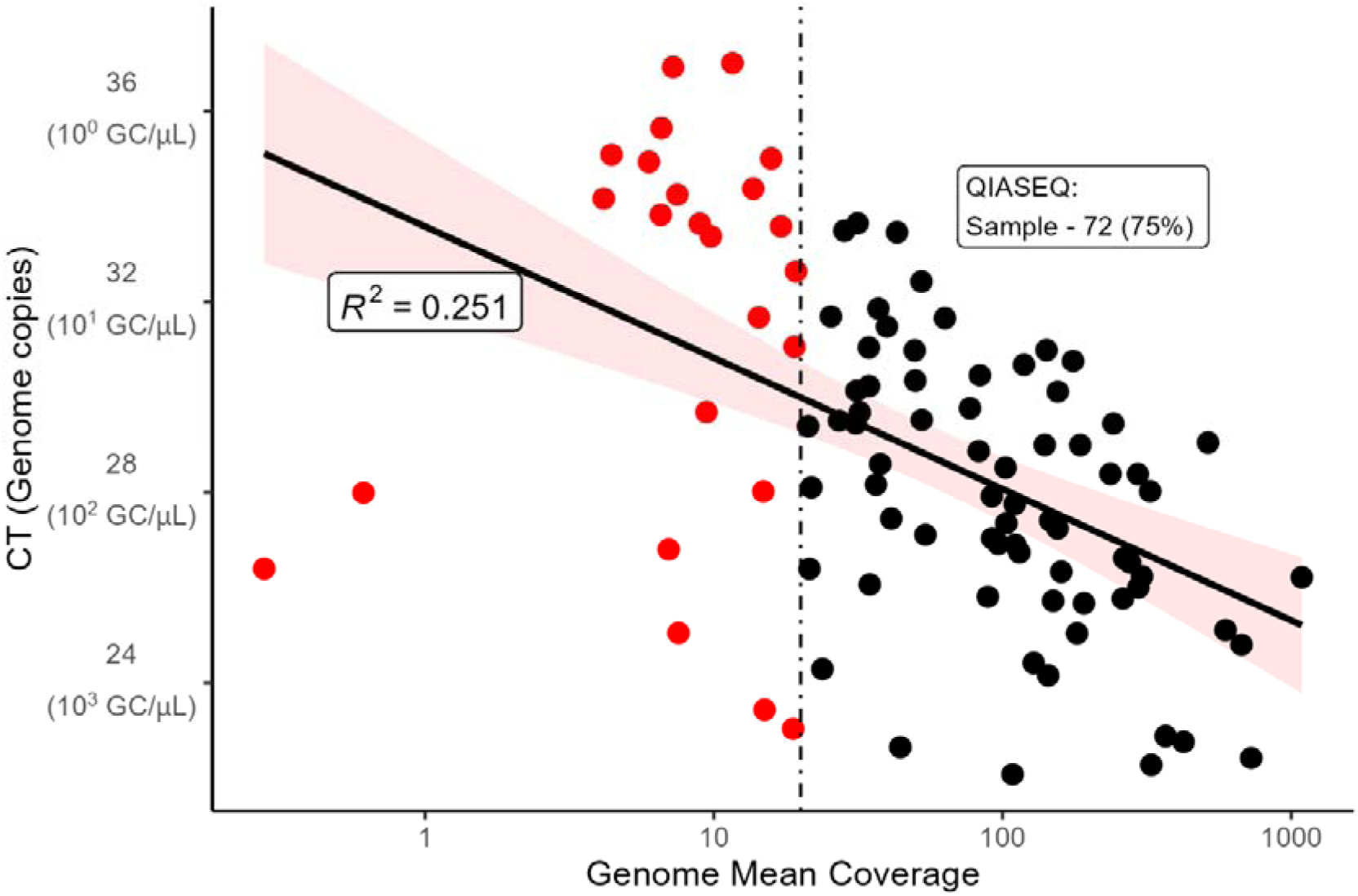
Correlation between QIAseq xHYB MTB data depth coverage and Ct values for 96 MTBC sediments analyzed with dWGS. The coefficient of determination (R^2^) represents the proportion of variance from the linear interpolation.

### Genotyping

Deeplex Myc-TB XL lineage predictions, which rely on spoligotyping and phylogenetic variants in target genes, showed 94.2% concordance at the lineage (L) level (81/86) and 93.0% concordance at the sub-lineage level (80/86) with cWGS genotyping (based on genome-wide SNP profiling by MTBseq pipeline(29)) (**Additional file 2**). Among six discordant samples, one was assigned to L4.7 by cWGS but classified as “L5.6 (*M. africanum*), animal strains” by tNGS (lineage-level discrepancy). Interestingly, the same sample results misclassified also by dWGS, as L4.3. The remaining five discrepancies involved samples identified as L4.8 by cWGS, of which four were reported as “not applicable” and one as L4.3 by tNGS (sub-lineage discrepancy). A mixed infection of MTBC and a nontuberculous mycobacterium (NTM) (i.e., *Mycobacterium shinjukuense,* 10.8%) was detected by tNGS in one sample.

Comparison of lineage assignments between cWGS and dWGS, both based on genome-wide SNP profiling (MTBseq (29) and CLC Genomics Workbench, respectively), revealed 100% concordance at the lineage level. Considering sub-lineage assignments, concordance was 97.1% (67/69 MTBC clinical sediments). The two discordant samples differed only at the sublineage level: one was classified as Manu2 by dWGS but as L4.1.1.3 Euro-American (X-type) by cWGS, and another (also misclassified by tNGS as L5.6) as L4.3 Euro-American (LAM5) by dWGS but as L4.7 Euro-American (mainly T) by cWGS.

### Drug Resistance detection

tNGS-based drug resistance detection, when benchmarked against cWGS on 86 MTBC clinical isolates successfully sequenced and analysed (**Table 1; Additional file 3**), demonstrated 100% sensitivity for all anti-TB drugs evaluated (BDQ, CFZ, DLM/Pa, EMB, ETH, INH, LEV, MXF, PZA, RIF, and STM), as well as specificity with minor specificity deviations observed for ETH (97%, 95% CI: 90–100) and INH (98%, 95% CI: 92–100). Indeed, two samples showed discrepancies in INH/ETH susceptibility results. Specifically, SNPs in the *fabG1* promoter region was detected exclusively by the Deeplex Myc-TB XL assay at allele frequencies of 40.4% and 49%, respectively. These variants were not confirmed by cWGS, while dWGS failed for the corresponding samples. The first sample carried, in addition to the *fabG1* loss of function, a *katG* S315T mutation (65.1%), confirmed by cWGS, and consistent with the INH-resistant phenotype. The second sample harboured only the *fabG1* promoter mutation associated with INH/ETH resistance, while phenotypic DST from culture indicated isoniazid susceptibility.

**Table 1:**
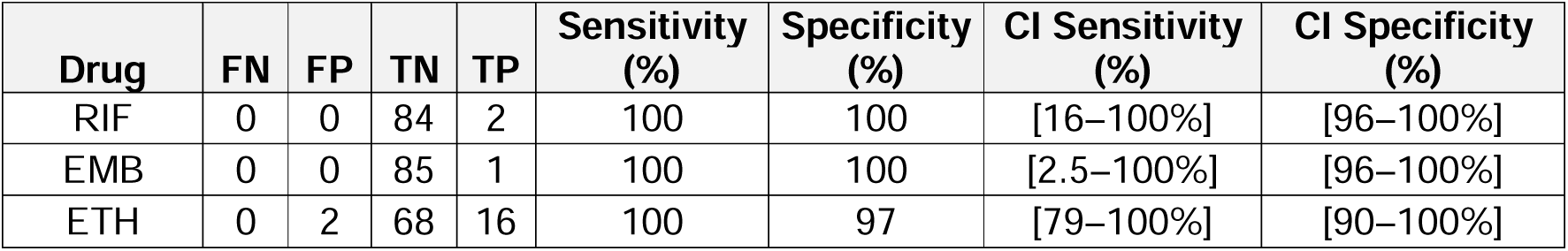
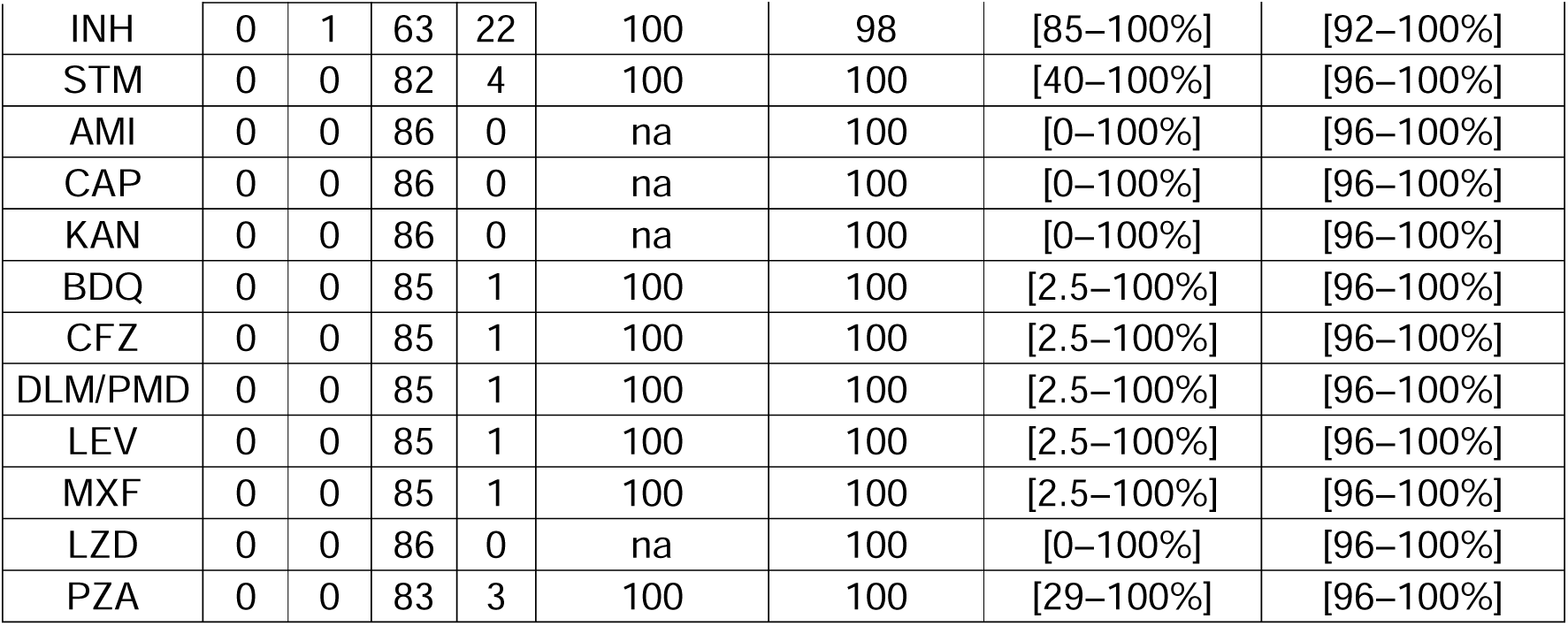
Performance of the Deeplex Myc-TB XL tNGS assay for detection of drug resistance in 86 MTBC clinical isolates compared to cWGS. Sensitivity, specificity, and corresponding 95% confidence intervals (CI) are reported for each drug. Test sensitivity could not be assessed (na) for AMI, KAN, CAP and LZD, as no known resistance-associated mutations were present in cWGS for these drugs. FN: False Negative; FP: False Positive; TN: True Negative; TP: True Positive.

| Drug | FN | FP | TN | TP | Sensitivity (%) | Specificity (%) | CI Sensitivity (%) | CI Specificity (%) |
| --- | --- | --- | --- | --- | --- | --- | --- | --- |
| RIF | 0 | 0 | 84 | 2 | 100 | 100 | [16–100%] | [96–100%] |
| EMB | 0 | 0 | 85 | 1 | 100 | 100 | [2.5–100%] | [96–100%] |
| ETH | 0 | 2 | 68 | 16 | 100 | 97 | [79–100%] | [90–100%] |
| INH | 0 | 1 | 63 | 22 | 100 | 98 | [85–100%] | [92–100%] |
| STM | 0 | 0 | 82 | 4 | 100 | 100 | [40–100%] | [96–100%] |
| AMI | 0 | 0 | 86 | 0 | na | 100 | [0–100%] | [96–100%] |
| CAP | 0 | 0 | 86 | 0 | na | 100 | [0–100%] | [96–100%] |
| KAN | 0 | 0 | 86 | 0 | na | 100 | [0–100%] | [96–100%] |
| BDQ | 0 | 0 | 85 | 1 | 100 | 100 | [2.5–100%] | [96–100%] |
| CFZ | 0 | 0 | 85 | 1 | 100 | 100 | [2.5–100%] | [96–100%] |
| DLM/PMD | 0 | 0 | 85 | 1 | 100 | 100 | [2.5–100%] | [96–100%] |
| LEV | 0 | 0 | 85 | 1 | 100 | 100 | [2.5–100%] | [96–100%] |
| MXF | 0 | 0 | 85 | 1 | 100 | 100 | [2.5–100%] | [96–100%] |
| LZD | 0 | 0 | 86 | 0 | na | 100 | [0–100%] | [96–100%] |
| PZA | 0 | 0 | 83 | 3 | 100 | 100 | [29–100%] | [96–100%] |

As regard dWGS-based drug resistance detection, it was benchmarked against cWGS for 69 MTBC clinical isolates successfully sequenced and analysed, **Table 2; Additional file 3**).

**Table 2:**
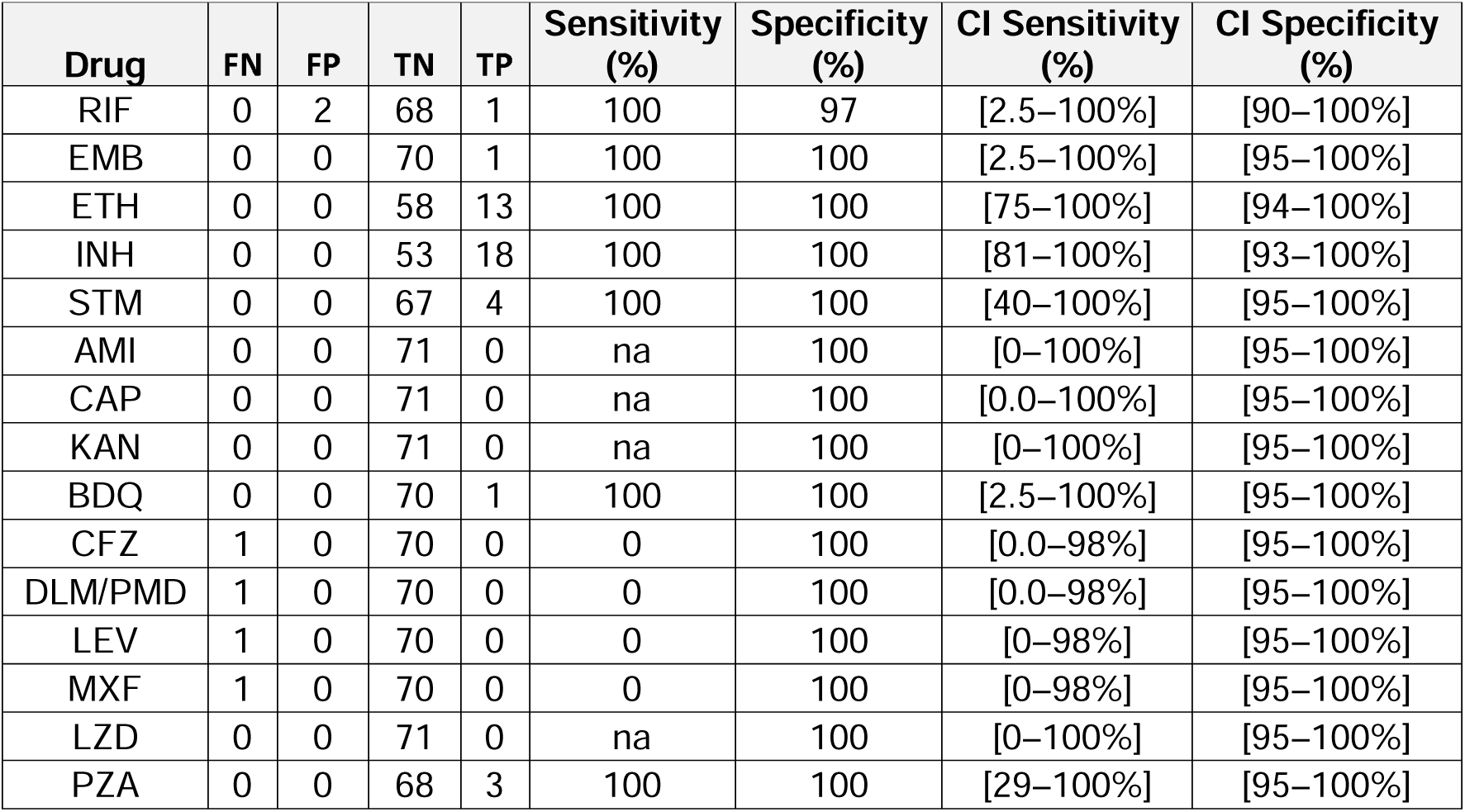
Performance of the QIAseq xHYB MTB direct WGS assay for drug resistance detection in 69 MTBC clinical isolates compared to cWGS. Sensitivity, specificity, and 95% confidence intervals (CI) are shown for each drug. Test sensitivity could not be assessed (na) for AMI, KAN, CAP and LZD, as no known resistance-associated mutations were present in cWGS for these drugs. FN: False Negative; FP: False Positive; TN: True Negative; TP: True Positive.

Sensitivity reached 100% for most first- and second-line anti-TB drugs (BDQ, EMB, ETH, INH, PZA, RIF, and STM), while five resistant cases were missed or misinterpreted for DLM, LEV/MXF and CFZ. Specificity remained 100% for all drug evaluated, with slight deviation observed for RIF (97%, 95% CI: 90–100).

Of the five discrepant isolates, two harboured *rpoB* Ser450Phe mutations detected exclusively by dWGS at variant frequencies of 28% and 61%, which were not identified by cWGS and tNGS, and accounted for the drop in RIF specificity to 97.1%. The remaining discrepancies were attributable to differences in mutation interpretation rules: one sample carried a *fbiC* loss of function insertion associated with DLM resistance, identified by cWGS (100%) and tNGS (86%) but not reported as resistance-associated by dWGS software (although called). Similarly, in another sample, a Rv0678 loss of function insertion associated to CFZ resistance was called but not interpreted by the dWGS software. Lastly, one sample exhibited a *gyrA* Asp89Asn mutation linked to fluoroquinolone resistance, detected at 22% and 21% allele frequencies by cWGS and tNGS, respectively, but not called in the dWGS analysis at the set calling thresholds.

### Transmission analysis

Transmission analysis of the 62 MTBC samples sequenced at suitable quality by both cWGS and dWGS confirmed the presence of a single cluster comprising two samples. This cluster was consistently identified in the minimum spanning trees (MSTs) generated from both cWGS- and dWGS-derived cgMLST profiles, indicating concordant results between the two sequencing approaches (**Additional file 4 - Suppl. Fig. 3A, B**). No additional clusters were observed among the remaining samples.

## Discussion

Our study evaluated for the first time two culture-free sequencing approaches, namely the Deeplex Myc-TB XL for tNGS and QIAseq xHYB MTB for dWGS, on a panel of MTBC positive decontaminated sediments (23). The performance of these workflows was evaluated for their qPCR-based limit of detection and, when benchmarked against cWGS, for drug resistance prediction accuracy, genotyping concordance, and suitability for transmission analysis.

The QIAseq xHYB MTB panel enrichment used for dWGS targets the entire genome of all seven major MTBC lineages, tolerating mismatches for unannotated or emerging variants (22). The Deeplex Myc-TB XL assay for tNGS builds upon the WHO-endorsed Deeplex Myc-TB kit (13), expanding the panel from 18 to 29 targets, including the drugs recommended for RR/MDR-TB treatment like bedaquiline and pretomanid. Both tNGS and dWGS assays design align with current WHO MTBC mutation catalogue (14) and treatment guidelines (2), supporting standardized interpretation of sequencing results.

The extracted DNA dataset, quantified using an in-house validated MTBC-specific qPCR assay targeting a single-copy genomic locus (26), encompassed MTBC genome copy concentrations ranging from 1 to 10^4^ gc/µL (**Additional file 4 - Suppl. Fig. 1**). This broad range of bacterial loads in the dataset enabled us to investigate performance across paucibacillary and high-burden samples and to stratify analyses according to loads. Overall, tNGS demonstrated high analytical robustness and accuracy. Interpretable sequencing results were obtained for 96.6% of samples, reporting a stronger correlation with Ct values than dWGS. The overall association was moderate (R² = 0.4), but the apparent correlation was weakened by a plateau effect at higher bacillary loads.

tNGS demonstrated a lower detection limit (∼10 genome copies total), outperforming both dWGS (∼100 genome copies total) and the previous Deeplex Myc-TB version, which required 10^2^–10^3^ genome copies for successful analysis (13).

dWGS produced sequencing data suitable for drug resistance analysis in roughly three-quarters of samples from trial participant. The correlation between Ct values and sequencing coverage was weaker than for tNGS, suggesting greater sensitivity to the DNA extraction method, DNA quality, and library preparation workflow. Unlike tNGS, which targets a limited genomic region (∼60 kb) and therefore achieves high depth even from low-input samples, dWGS distributes reads across the whole genome and lacks a targeted amplification step, resulting in lower per-locus coverage at comparable input levels.

Our findings provide important new evidence advancing the feasibility and performance of MTBC culture-free WGS. Earlier studies demonstrated that direct sequencing from clinical specimens is possible but faced major limitations. Doughty et al. first showed that dWGS could be performed directly from specimens, but extremely low genome coverage (0.002–0.7×) and overwhelming human DNA contamination (22-99% of reads) prevented meaningful genomic interpretation (18). Subsequent enrichment-based strategies, including host DNA depletion and hybrid capture with biotinylated probes, significantly improved performance but introduced workflow complexity and still yielded modest coverage (17, 19–21). Goig *et al.* achieved reliable resistance prediction and, for the first time, resolved transmission clusters using RNA bait capture technology (20), but even with enrichment, mean coverage typically ranged from 12–30x (20). In contrast, our study demonstrates that hybrid-capture dWGS using the QIAseq xHYB MTB panel can achieve substantially higher mean genome depths (150×) directly from clinical sputum sediments, a several-fold improvement over previously reported direct or enriched approaches. This level of coverage enables more confident variant calling, more robust resistance prediction, and greater suitability for high-resolution phylogenetic analyses than prior methods. Notably, we show that such deep coverage is achievable when hybrid capture is combined with efficient host DNA depletion using the Molzym extraction workflow, highlighting the importance of integrating optimized upstream sample-processing steps. Earlier versions of this kit have already been successfully applied to culture-free dWGS of MTBC in previous studies (17). Together, these results demonstrate that high-depth, culture-free WGS of MTBC is not only technically feasible but can meet the performance requirements for clinically meaningful genomic analyses, representing a significant advance over earlier dWGS protocols.

A bacillary-load–based strategy to select between tNGS, dWGS, and, when required, cWGS could improve both efficiency and reliability of sequencing for DR-TB diagnosis. The original Xpert scores were not recorded in the trial; bacillary load was therefore quantified in this study by qPCR, enabling inference of corresponding Xpert categories. Based on our findings, samples with fewer than 10–50 genome copies (approximately corresponding to Xpert Ultra trace scores) may require culture prior to sequencing, although tNGS may still be attempted. Samples with Xpert values categorized as very low or up to ∼10² genome copies are generally suitable for tNGS but unlikely to yield successful dWGS. In contrast, samples with Xpert low, medium, or high categories, corresponding to ≥10³ genome copies, are appropriate for either tNGS or dWGS, depending on the desired genomic resolution and turnaround time (**Additional file 4** - **Supp. Fig. 4**).

Our findings highlight important differences in the performance and analytical behavior of tNGS and dWGS for MTBC drug resistance profiling. As expected for an amplicon-based assay, tNGS demonstrated very high diagnostic accuracy, with near-perfect sensitivity and specificity across drugs. The two INH/ETH discrepancies identified solely by tNGS likely reflect unfixed variants present in the original specimen but lost during culture, consistent with previous evidence that cWGS can fail to capture within-host heterogeneity (30–32). This suggests that tNGS may offer an advantage in detecting emerging or minority variants that could be clinically relevant in early resistance development.

dWGS also performed strongly overall; however, its reduced sensitivity and two false-positive signals for rifampicin underscore some inherent challenges of applying culture-free WGS directly to sputum. For instance, background noise in conserved regions like *rpoB* and *rrs* suggests that hybrid-capture enrichment can inadvertently co-capture non-MTBC DNA, making the low-frequency artifacts more likely a consequence of the capture method than of the analysis software, as supported by the detection of the same variants in alternative pipelines. Given such interference, we applied stringent filters beyond the default analysis settings to minimize false positives and limit outputs to high-confidence variants. The misclassification of variants in *fbiC* and Rv0678 further suggests that software implementations of the WHO mutation catalogue may still require refinement, especially for genes where multiple rules or variant categories apply. Additionally, a *gyrA* Asp89Asn mutation reported by cWGS (22%) was not identified in the dWGS call set, although detected at 16% using an alternative pipeline (MTBseq), suggesting that low-frequency variants near analytical thresholds may be missed due to limited depth and pipeline-specific filtering. Together, these observations emphasize that while dWGS is a powerful method for comprehensive and rapid resistance prediction, its reliability is dependent on both bacterial load and the degree of off-target capture. Improving host-depletion efficiency, refining probe design, and optimizing variant-calling thresholds will be essential to enhance robustness, particularly for regions prone to cross-species interference. Furthermore, the results highlight the need for standardized, WHO-aligned and validated bioinformatic workflows to ensure consistent and accurate resistance prediction across sequencing platforms.

Interestingly, the *rpoB* Ile491Phe mutation, typically missed by Xpert and other rapid assays (33), was successfully detected by both tNGS and dWGS, underscoring the enhanced resolution of NGS-based approaches for comprehensive resistance profiling.

Lineage assignment from both culture-free sequencing approaches showed high overall concordance with cWGS, confirming the reliability of direct genotyping from clinical sediments. Agreement was complete for dWGS at the lineage level, while tNGS showed few discrepancies, including one misclassified lineage and several borderline sublineage assignments. Such assignment likely reflects the insufficient typing resolution power of genomic regions targeted by the assay, highlighting the inherent limitation of tNGS to predefined loci versus WGS for lineage determination and comprehensive genotyping. A single mixed MTBC–NTM signal detected by tNGS was not confirmed by cWGS, likely reflecting low-abundance NTM DNA not captured after culture step or software misclassification.

High-quality sequencing data suitable for transmission analysis were obtained for 64.6% of dWGS samples, demonstrating that this approach can resolve MTBC epidemiological patterns without the need for prior culture. Despite limited cluster representation due to random multi-site sampling, dWGS reliably reproduced cWGS-inferred genotyping and transmission networks when adequate coverage breadth was achieved. This function is highly relevant for guiding public-health interventions in both high-burden and low-incidence settings, including for rapid tracking of cross-border transmission in TB elimination contexts such as Europe (4). dWGS can also be valuable in clinical trials, where participants typically present with higher bacillary loads. It supports endpoint resolution (e.g., culture-independent discrimination of relapse vs reinfection), enables detection of preexisting or emergent drug resistance, and permits evaluation of intervention performance across diverse pathogen genomic backgrounds, including MTBC lineages.

The dWGS workflow exhibited greater procedural complexity than tNGS, leading to longer turnaround times and higher costs, consistent with its whole-genome coverage compared with the targeted scope of tNGS. The QIAseq xHYB MTB assay requires qPCR quantification of samples for pooling before hybrid capture and library preparation, which lengthens the overall workflow. In our study, this resulted in approximately five days to process 96 samples. The practicality of this pooling strategy under routine clinical conditions has yet to be determined. By contrast, the Deeplex Myc-TB XL protocol can be completed in under three days, thanks to a streamlined workflow that eliminates post-PCR clean-up and quantification steps (unlike the previous assay version), as amplicons are processed directly for Illumina library preparation. Future efforts should focus on developing more cost-effective, streamlined, and automation-compatible dWGS enrichment strategies that can enable scalable sequencing for tuberculosis diagnostics and surveillance at sustainable costs.

This study has some limitations. The sample collection was not enriched for DR MTBC strains, as per trial design, and resistant isolates were unavailable for certain drugs. Consequently, confidence intervals for some sensitivity and specificity estimates are wide, reflecting the limited number of isolates in specific resistance subgroups and reducing the precision of these estimates. These factors indicate that our findings should be validated in larger, more diverse cohorts. In addition, the two evaluated kits were either still in beta testing or had only recently been released, meaning that workflow conditions and analysis-software interpretation rules may evolve with future product updates.

## Conclusions

This study provides the first head-to-head evaluation of two next-generation, culture-free sequencing strategies for direct detection of MTBC from sputum sediments. Both approaches advance current diagnostic capabilities by enabling comprehensive drug resistance profiling without culture, including detection of resistance markers for newly introduced agents such as bedaquiline and pretomanid. The tNGS assay delivered enhanced sensitivity and coverage of an expanded resistance panel, whereas the dWGS workflow enabled genome-wide interrogation of resistance loci and high-resolution transmission inference. Together, these findings support the clinical, public-health and trial utility of culture-free sequencing as a transformative tool for RR/MDR-TB management, surveillance, and evaluation of new drugs and vaccines.

## Data Availability

All data generated or analysed during this study are included in this published article, and its supplementary information files.

## List of abbreviations

WHO: World Health Organization
DR-TB: drug-resistant tuberculosis
RR/MDR-TB: rifampicin-resistant/multi-drug-resistant tuberculosis
BPaLM: bedaquiline, pretomanid, linezolid, moxifloxacin regimen
NGS: next-generation sequencing
MTBC: Mycobacterium tuberculosis complex
tNGS: targeted next-generation sequencing
WGS: whole-genome sequencing
dWGS: direct whole-genome sequencing
cWGS: culture-based whole-genome sequencing
pDST: phenotypic drug susceptibility testing
qPCR: quantitative real-time polymerase chain reaction
Ct: cycle threshold
gc: genome copies
INH: isoniazid
PZA: pyrazinamide
EMB: ethambutol
FQ: fluoroquinolones
BDQ: bedaquiline
LZD: linezolid
CFZ: clofazimine
AMK: amikacin
STM: streptomycin
KAN: kanamycin
CAP: capreomycin
PMD: pretomanid
ETH: ethionamide
LEV: levofloxacin
MOX: moxifloxacin
DLM: delamanid
TP: true positives
FN: false negatives
TN: true negatives
FP: false positives
cgMLST: core genome multi-locus sequence typing
MST: minimum spanning tree
L: lineage
SNP: single nucleotide polymorphism
NTM: non-tuberculous mycobacteria

## Declarations

## Ethics approval and consent to participate

The trial was done in accordance with the Declaration of Helsinki, the International Council for Harmonisation Guideline for Good Clinical Practice (version E6, revision 2), and national Good Clinical Practice guidelines, and is registered with ClinicalTrials.gov, NCT03512249 (23). The study protocol was approved by the South Africa Health Products Regulatory Authority (Pretoria, South Africa; approval number 20180624), by the Tanzania Medicines and Medical Devices Authority (Dar es Salaam, Tanzania; approval number TFDA0019/CTR/00147/18), and by all relevant institutional ethics committees in South Africa and Tanzania (23).

All participants consented to participate in the trial and to the sample storage for biobank and TB research (23).

## Consent for publication

Not applicable.

## Availability of data and materials

dWGS and tNGS data generated in this study are available in the Sequence Read Archive of the National Center for Biotechnology Information as raw FASTQ files, under study accession number PRJNA1392017 (27). The raw FASTQ files from cWGS data have been previously deposited in the NCBI Sequence Read Archive under the study accession number PRJNA1171160 (24). All data generated or analysed during this study are included in this published article, and its supplementary information files.

## Funding

The study was supported by the EU through The European and Developing Countries Clinical Trials Partnership (EDCTP2) (grant number RIA2016V-1631, POR TB consortium).

GenoScreen provided the kit and analysis software for free. Qiagen provided the analysis software for free. The two companies had no role in study design; data collection, analysis and interpretation; decision to publish, or preparation of the manuscript.

## Authors’ contributions

II: design of the work; acquisition, analysis and interpretation of data; writing of the original draft.

FDM: analysis and interpretation of data; substantial review of the work.

KM: acquisition and analysis of data.

PM: interpretation of data; substantial review of the work.

PORTB: substantial review of the work.

AMC: conception and design of the work; analysis and interpretation of data; writing of the original draft; supervision.

DMC: conception and design of the work; interpretation of data; substantial review of the work; supervision.

All authors read and approved the submitted version of the manuscript.

## Acknowledgements

This work represents the outcome of experimental research conducted as part of the PhD thesis of Ilaria Iannucci, PhD student at Università Vita-Salute San Raffaele, affiliated with the Emerging Bacterial Pathogens Unit, Division of Immunology, Transplantation and Infectious Diseases, IRCCS San Raffaele Scientific Institute.

## *POR TB study group* collaborating authors

Álvaro H Borges, Statens Serum Institut, Copenhagen, Denmark

Andrea M Cabibbe, IRCCS San Raffaele Scientific Institute, Milano, Italy

Gavin Churchyard, The Aurum Institute, Johannesburg, South Africa; University of Witwatersrand, Johannesburg, South Africa; Vanderbilt University, Nashville, TN, USA

Daniela M Cirillo, IRCCS San Raffaele Scientific Institute, Milano, Italy

Rodney Dawson, University of Cape Town Lung Institute, Cape Town, South Africa

Andreas H Diacon, TASK, Cape Town, South Africa

Mark Haterill, University of Cape Town, Cape Town, South Africa

Rasmus Mortensen, Statens Serum Institut, Copenhagen, Denmark

Elisa Nemes, University of Cape Town, Cape Town, South Africa

Marisa Russell, IAVI, Cape Town, South Africa

Issa Sabi, National Institute for Medical Research, Mbeya, Tanzania

Thomas Scriba, University of Cape Town, Cape Town, South Africa

Per Skallerup, Statens Serum Institut, Copenhagen, Denmark

Elana Van Brakel, IAVI, Cape Town, South Africa

