## Additional file 4 for "Towards Culture-Free Sequencing of *Mycobacterium tuberculosis*: Evaluating New Targeted and Whole-Genome Approaches for Genotyping and Drug Resistance Profiling"

**Supplementary Figure 1: A, B.** qPCR quantification of MTBC genome copies (gc/µL): distribution of samples by concentration range

(A)	(B)

gc/µL dataset distribution

N° samples

| MTB gc/µL | N° samples |
| --- | --- |
| 1-10 | 21 |
| 10-100 | 34 |
| 100-1000 | 29 |
| >1000 | 12 |

| **genome copies (gc)/µL** | **Cycles threshold (Ct) range** | **gc/µL mean** | **N samples** | **%** |
| --- | --- | --- | --- | --- |
| **<10** | 31-37 | 2.6 | 21 | 21.9 |
| **10-100** | 28-31 | 40.7 | 34 | 35.4 |
| **100-1000** | 24-27 | 330 | 29 | 30.2 |
| **>1000** | <24 | 4012 | 12 | 12.5 |

**Supplementary Figure 2**: **A**. Flowchart indicating the number of MTBC clinical sediments selected, analysed and interpreted for each study; **B.** Flowchart representing the method workflow for each sample analysed, including WGS from culture, tNGS (Deeplex XL Myc-TB) and dWGS (QIAseq xHYB MTB).

(2A)

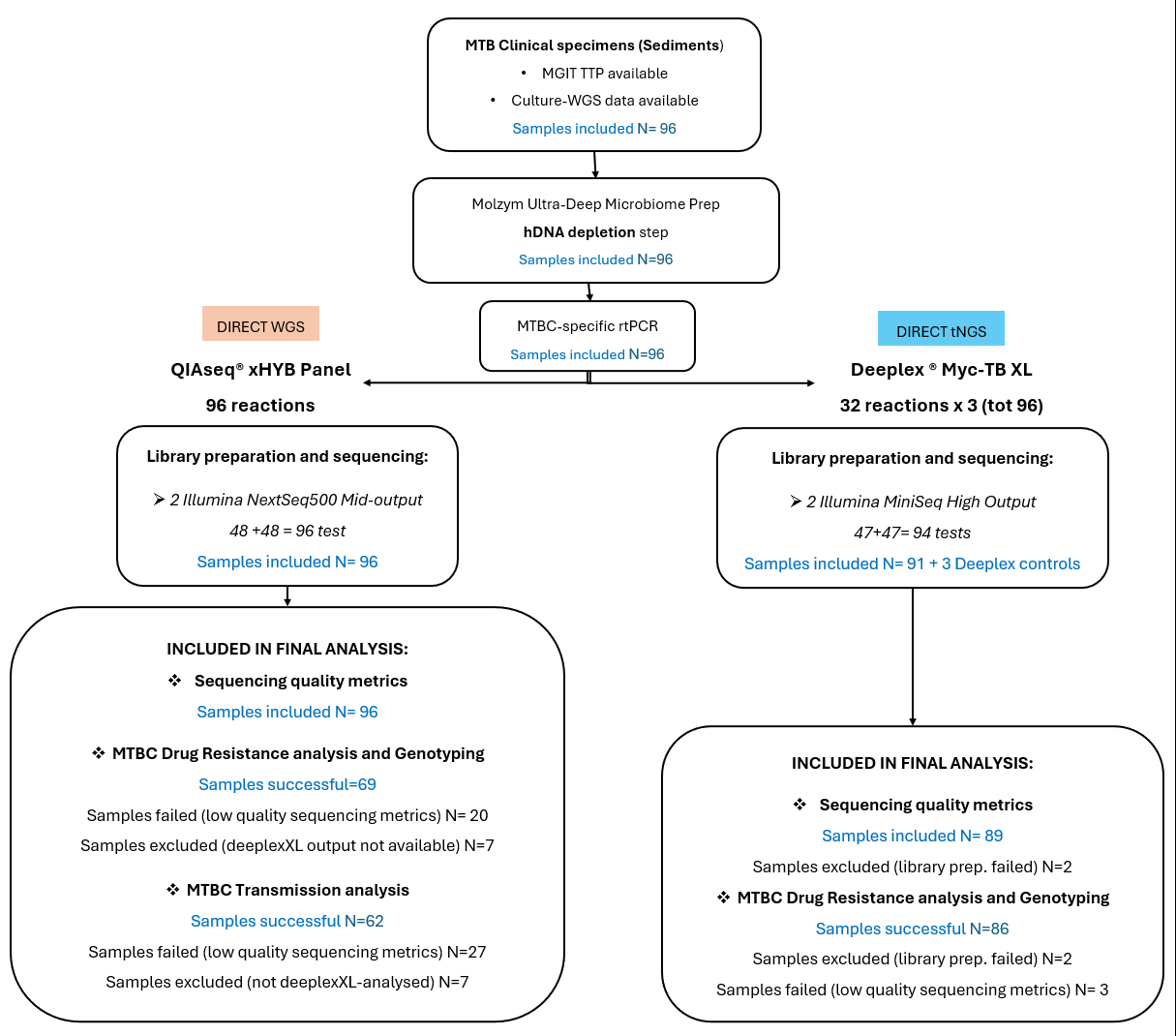

(2B)

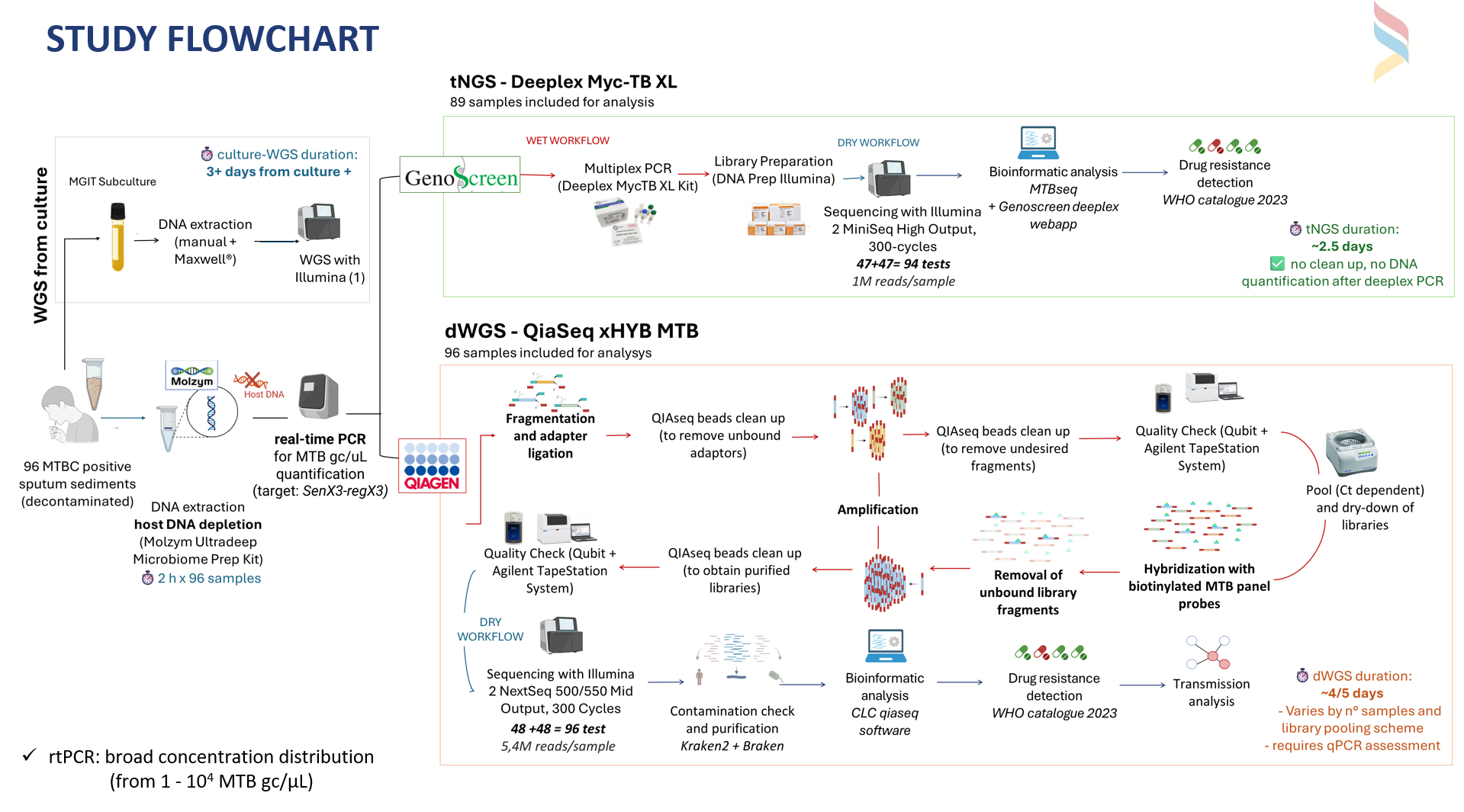

**Supplementary Figure 3:** Transmission Analysis **A.** MST illustrating clusters from dWGS, with MST cluster 1 highlighted in pink**; B.** MST illustrating clusters from cWGS, with the same MST cluster 1 highlighted in red. Cluster distance threshold = 5 alleles. Generated with core genome multi-locus sequence typing (cgMLST) scheme implemented in *Ridom SeqSphere+®* software.

(3A)

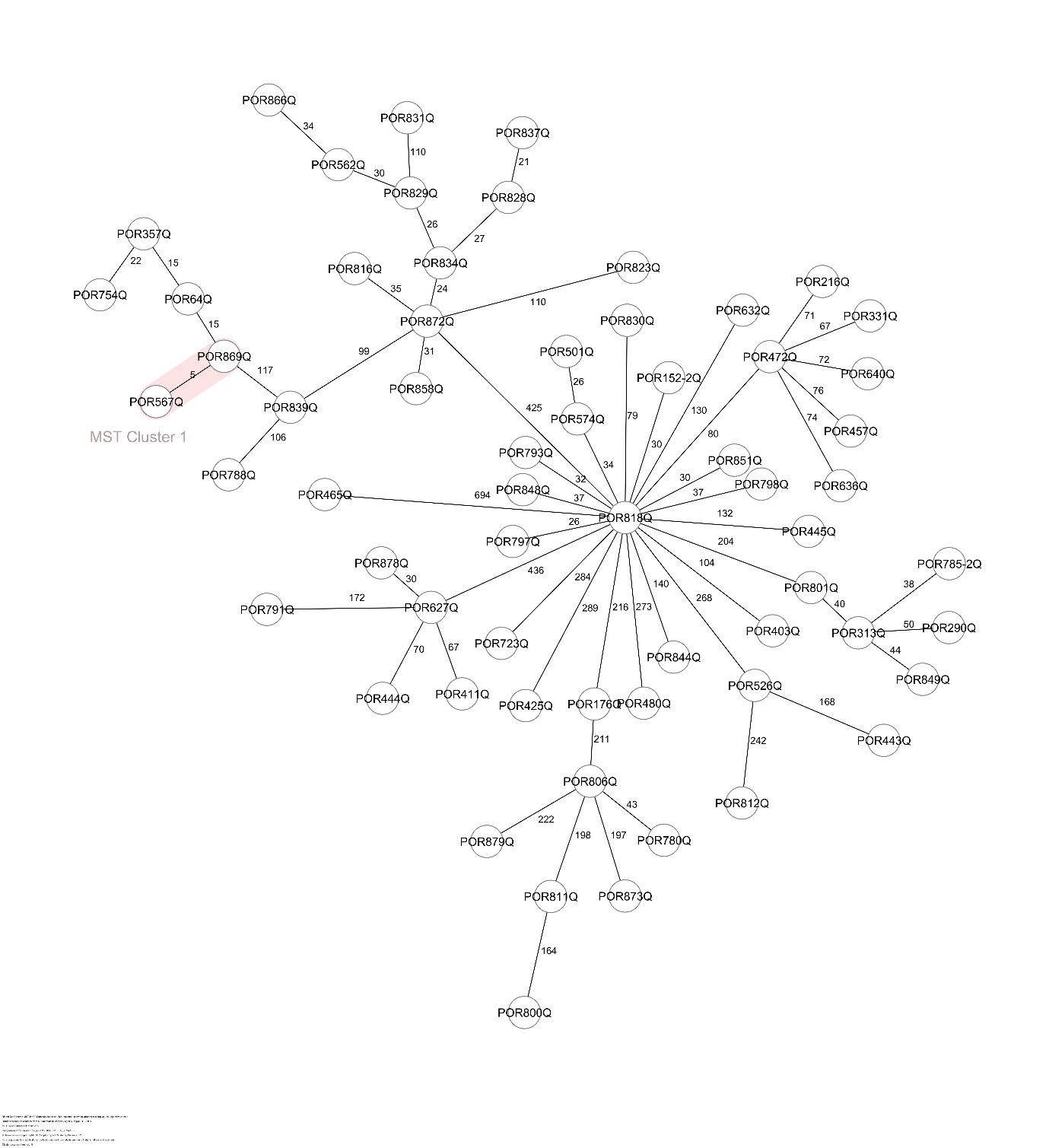

(3B)

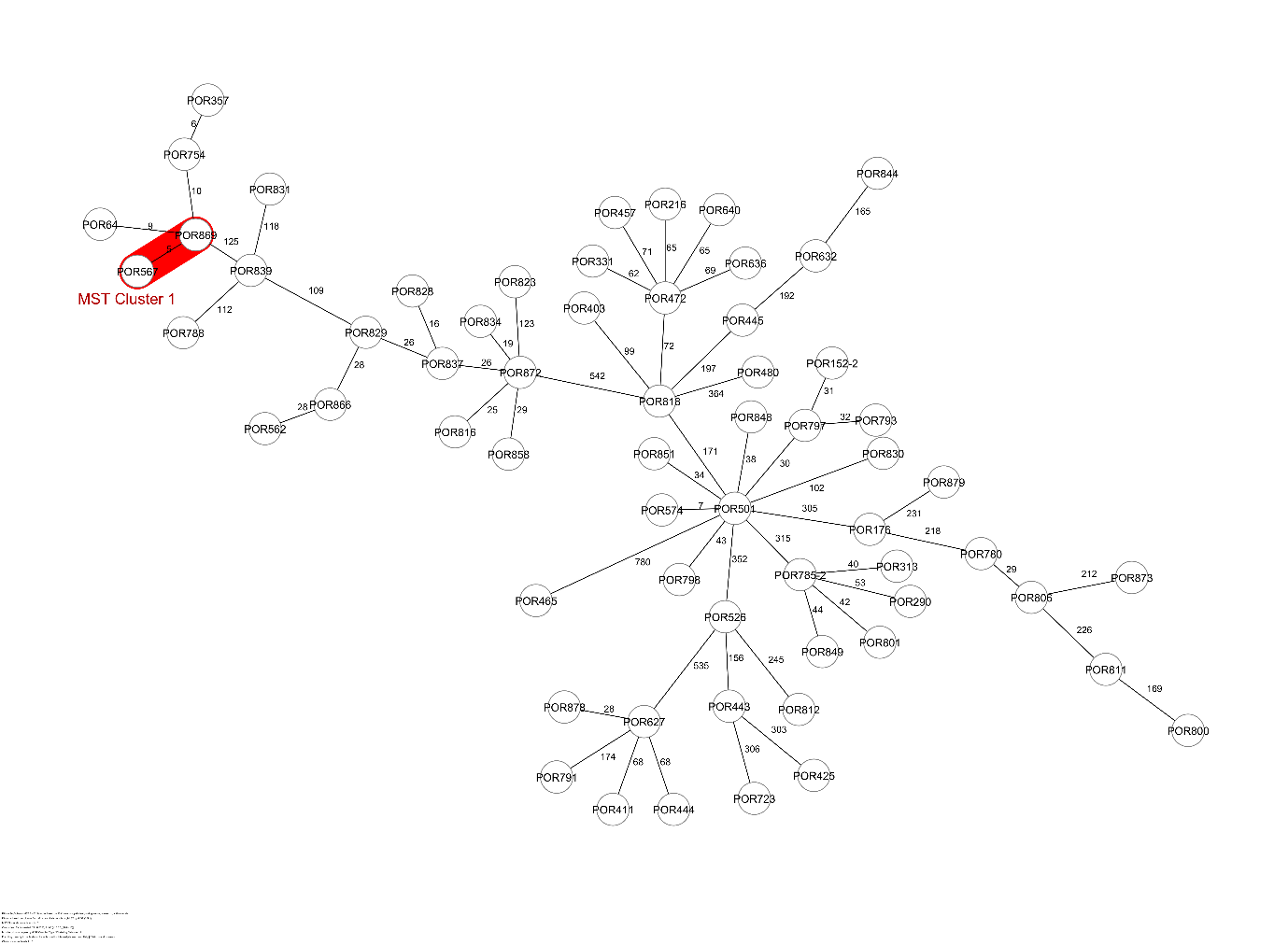

**Supplementary Figure 4:** Decision algorithm for implementation**,** bacillary-load–guided sequencing strategy for DR-TB diagnosis

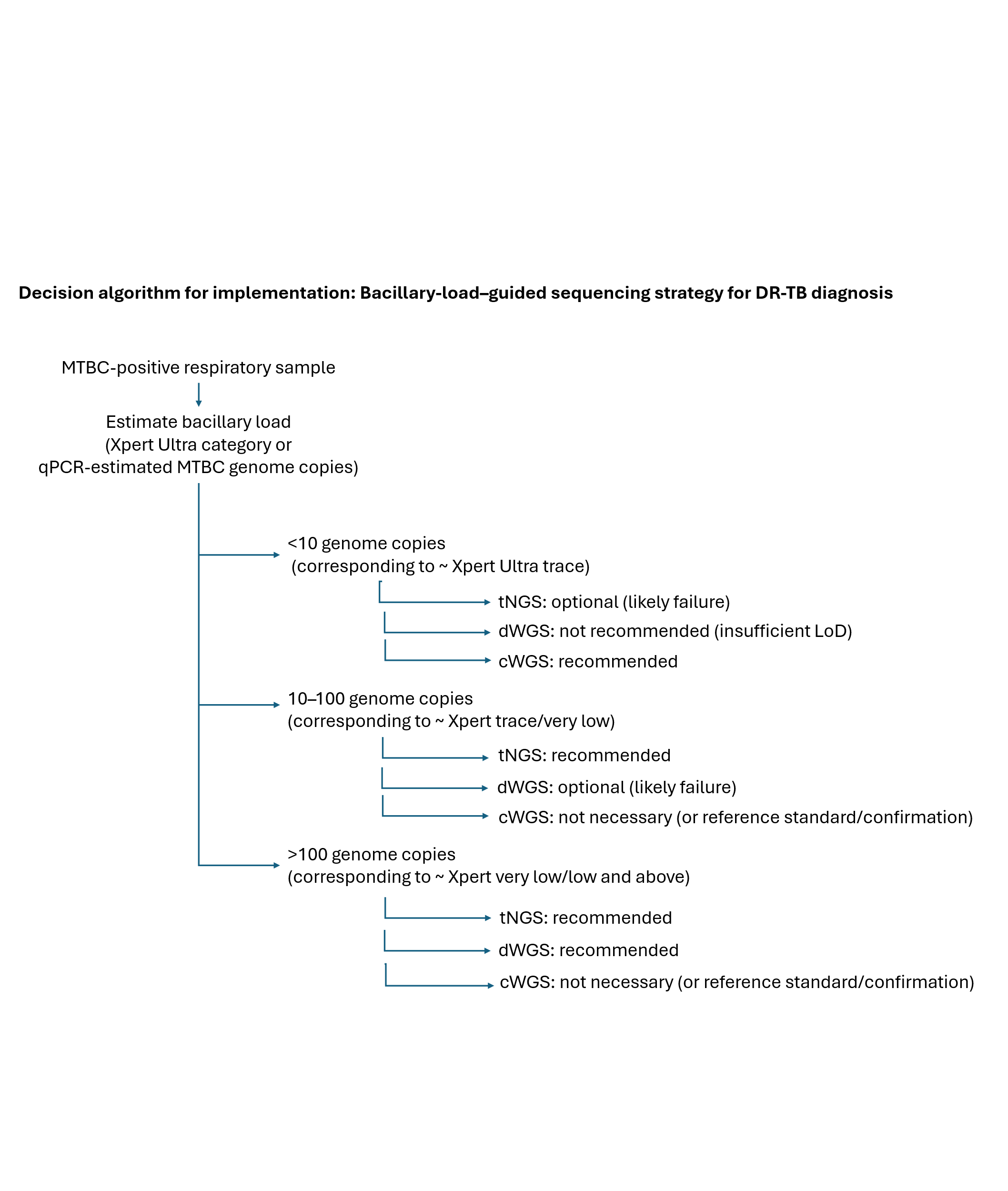
